# Assessing dengue knowledge, attitudes and preventive practices using participatory surveillance cohorts

**DOI:** 10.1101/2025.11.05.25339567

**Authors:** Lucille Calmon, Alessandro de Gaetano, Mattia Mazzoli, Nicolò Gozzi, Marion Debin, Clément Turbelin, Romain Marmorat, Nicola Perra, Alain Barrat, Vittoria Colizza, Daniela Paolotti

## Abstract

**Background:** The dengue virus, transmitted by *Aedes* mosquitoes, causes an estimated 100–390 million infections annually. With no treatment available, mitigation and prevention involves reducing mosquito breeding and limiting feeding opportunities. Due to changing climate conditions, *Aedes* mosquitoes are increasingly established in Europe causing more local cases. In this context, understanding population knowledge, attitudes, and practices in areas where dengue is an emerging threat is necessary for effective prevention.

**Methods:** We designed a survey collecting knowledge, attitudes, and beliefs about dengue in the general population, leveraging the French and Italian 2024 cohorts of the participatory surveillance network InfluenzaNet. Responses from 2,500 participants in mainland France and 404 in Italy were pooled for the analysis. Via bias-adjusted (i.e., age, sex and education) logistic and ordinal regression, we examined determinants of awareness of dengue and knowledge of its characteristics, adoption of preventive measures, and vaccination willingness.

**Results:** Over 91% of included participants were aware of dengue circulating in Europe or the rest of the world, and 16% had sought information. Seeking information and concerns about dengue were significant determinants of knowledge, preventive measures adoption, and vaccination willingness. Better knowledge of dengue characteristics, vaccination status, residence in areas with detected autochthonous cases, and presence of comorbidities were associated with greater adoption of preventive measures. Living in rural areas was associated with lower knowledge and vaccination willingness but higher adoption of preventive measures. Finally, 0.4% of participants had received a dengue vaccine (not recommended to the general population), and 54.6% would accept a vaccine if offered. Low-risk perception was the most common reason for refusal.

**Conclusions:** Our study highlights the interplay between knowledge, attitudes, behaviours, socio-demographic, individual and contextual characteristics of the participants related to dengue in a non-endemic context. Information search and concern levels correlated with knowledge, adoption of measures, and willingness to vaccinate, while, knowledge was associated with the adoption of preventive measures, alongside perceived efficacy, local epidemiological context, occupation of participants, sex, vaccination and symptom history. Targeted communication strategies accounting for these factors are essential to enhance preparedness and outbreak prevention in regions where dengue represents an emerging threat.

## Background

The virus dengue^1^, transmitted to humans via the *Aedes aegypti* and *Aedes albopictus* mosquitoes, causes an estimated 100-390 million infections, and more than 40 thousands deaths worldwide each year^2–4^. While a vaccine granting moderate protection is recommended in regions with high dengue transmission^5,6^, there is currently no medical cure against dengue. Consequently, strategies to prevent and manage outbreaks heavily rely on vector control measures limiting mosquito proliferation and breeding, as well as feeding opportunities^7–10^. Community involvement in these strategies is paramount to maintain engagement with prevention programs^11,12^, which can present a challenge in newly affected regions where surveillance programs were only recently established^13^.

Dengue transmission has been historically concentrated in the tropics^14^, but regions previously spared are increasingly at risk due to changing climatic conditions^15–17^. Models accounting for different climate scenarios predict the future expansion of *Aedes* mosquitoes on the European continent^18,19^, possibly leading to sustained dengue transmissions as climatic conditions become better suited to the spread of the disease^20^. In Europe, autochthonous cases of dengue have been increasing since 2010^21,22^, especially in Italy, France, Spain and Greece. In 2024, an outbreak of at least 199 autochthonous cases was observed in Italy^23^, while 11 outbreaks totalling 83 autochthonous cases were detected in France^24^. In 2025, guidance from the ECDC on assessment and mitigation of *Aedes*-borne diseases anticipated an increase in the risk of local outbreaks of *Aedes*-borne in Europe^25^. This risk is particularly high for France, which faces high numbers of importations of dengue cases in mainland France from its overseas departments where dengue is endemic, including La Réunion and Les Antilles (Martinique and Guadeloupe). In the first quarter of 2024 for example, 82% of the 1679 dengue importations in mainland France came from Les Antilles^26^. In this context, the dengue vaccine Qdenga was approved in Europe in 2022, but is not recommended in mainland France. In the French overseas departments where dengue is circulating, it is recommended to children with a previous infection, and vulnerable adults. In Italy, it is recommended to travelers to endemic areas, for long (at least three weeks) or repeated trips, especially if the traveler has been previously infected with dengue. Promoting awareness and behavioural change to reduce the proliferation of mosquitoes and the transmission of dengue are pillars of public health strategies^27^. According to the health belief model, behavioural adaptations are generally shaped by the interplay of risk perception, benefits gained from the behavioural change, cues to actions and individual socio-demographic and psychological characteristics^28^. To effectively inform the population and encourage the adoption of preventive measures through tailored prevention programs, it is important to understand factors shaping knowledge, attitudes, prevention practices, and their interplay specific to mosquito-borne viruses and in particular dengue^29^. In endemic areas where the population is knowledgeable on mosquito-borne viruses characteristics and commonly adopt preventive measures^30–33^, these factors have been extensively investigated. Professional inactivity, lower education level, and older age were shown to be associated with worst knowledge of dengue characteristics in La Réunion^30^, while exposure to information on dengue, previous dengue infection and higher concern levels instead were associated with knowledge of dengue symptoms and transmission in Venezuela^31^. Knowledge about dengue itself was shown to drive the adoption of preventive practices in Venezuela^31^ and Malaysia^34^, but not in La Réunion^30^. The adoption of preventive practices was also more likely in women, individuals with higher education level, and those feeling to be frequently bitten by mosquitoes^30^, as well as for individuals reporting higher perceived susceptibility^31^ and perceived threat to society^32^.

In non-endemic regions, dengue specific knowledge, attitudes, preventive practices adoption and their drivers have been investigated in a few instances^35^. In Italy, Italians in the Lazio region were less knowledgeable on mosquito-borne viruses, and mosquito life cycles, compared to resident communities originally from the Indian subcontinent in 2016^36^ . Italians also reported fewer concerns for mosquito-borne viruses but higher annoyance with mosquitoes^36^. In French metropolitan departments colonised by *Aedes* mosquitoes, concern levels, annoyance with mosquitoes, perceived exposure to mosquito-borne diseases and perceived efficacy of preventive practices were associated with the adoption of preventive practices in 2016^37^, together with sex, education status, and previous identification of *Aedes albopictus* mosquitoes in 2012^38,39^. Longitudinal observations between 2012 and 2014 in a sample randomly selected in colonised departments found an increasing awareness of the mode of transmission of dengue^38^, but knowledge of other mosquito-borne viruses characteristics remained heterogeneous in both France^39^ and Italy^36^, while the adoption of preventive measures stayed moderate between 2012 and 2016^36,38,39^. The majority of studies in non-endemic regions however relied on small samples^35,40^, or focused on specific subpopulations, such as amateur gardeners^41^, medical professionals^42^ or residents in departments colonised by *Aedes* mosquitoes^37–39^. With the rapidly evolving establishment of *Aedes* vectors in Europe^43^, and the increasing detection of locally transmitted dengue cases^21^, it is necessary to monitor frequently the knowledge, attitudes and practices related to dengue in the general populations in Europe. This is especially important in the aftermath of the COVID-19 pandemic, which may have affected population awareness of health threats.

Participatory surveillance platforms^44^ like the European InfluenzaNet cohort network^45^ provide an efficient and low cost tool for this purpose, by leveraging volunteer participants continuously engaged with the platform. In parallel to the longitudinal respiratory syndromic surveillance conducted weekly, additional cross-sectional surveys can be rapidly deployed, as needed, e.g., to understand vaccination attitudes^46,47^ and behavioural changes^48^. This is of particular relevance when facing emerging threats where rapid insight is necessary, for example to capture risk perception at the start of the COVID-19 pandemic^49^.

In this study, we used the participatory cohorts of InfluenzaNet^45^ to survey populations on their knowledge, attitudes and practices in mainland France (including Corsica) and Italy, two territories increasingly exposed to dengue. The survey included questions on knowledge of dengue (i.e., its existence and main characteristics), information sources, perceived level of information, concern levels related to a set of consequences of a dengue outbreak, perceived efficacy of a set of preventive measures, adoption of these measures, attitudes towards vaccination, including reasons for refusal. We also enquired about possible recent dengue symptoms, and knowledge of a dengue case among relatives and acquaintances. A detailed profile for each participant including socio-demographic and health characteristics was also obtained from the InfluenzaNet platform. Using the gathered data, we conducted a statistical analysis to characterise factors associated with awareness of the existence of dengue, knowledge of its main characteristics, adoption of preventive measures and attitudes towards vaccination, in the scenario of a vaccine being offered. The findings of this study provide essential insights on knowledge, attitudes and practices in two European contexts, paving the way to the targeted design of interventions to raise awareness and encourage the adoption of preventive practices.

## Methods

### InfluenzaNet Data Collection

We collected data on knowledge, attitudes, and beliefs related to dengue fever among the French and Italian cohorts of InfluenzaNet, a survey network monitoring influenza-like illnesses across Europe^45^. Following an intake survey covering socio-demographic and health-related characteristics, enrolled participants are sent out weekly questionnaires on their influenza-like symptoms. Participation in weekly questionnaires is voluntary. For this study, participants were invited to complete dengue-specific questionnaires to assess their recent history of symptoms possibly linked to dengue, awareness of the disease, knowledge of key characteristics, concern levels, perception and adoption of preventive measures, and vaccination willingness, including reasons for refusal. The full survey text is available in the Supplementary Information (SI).

### Study Period and Inclusion Criteria

The data was collected between May and October 2024. In Italy, participants were invited to complete the questionnaire monthly between May 13, 2024, to October 27, 2024. These longitudinal data were collected to capture the impact of a potential summer outbreak, which will be the subject of a future study. For the present analysis, to avoid bias due to previous exposure to the questions, we included only the first complete submission of a questionnaire for each participant in Italy. In France, participants were invited to complete the questionnaire once on July 29, 2024, and we included responses filled out until August 6, 2024, after which daily numbers of responses significantly decreased. Participants were able to submit multiple responses within this single survey wave, as long as the questionnaire remained open. French responses were post processed to associate each participant with a single response (see SI). In both countries, we included participants who reported their sex, age and education level, provided that these met the age/sex/education census stratifications available for each country (see section *Post-Stratification*). We included residents of mainland France (including Corsica) and Italy. Included participants from the two countries were pooled for the analysis.

### Post-Stratification

The InfluenzaNet French and Italian samples differ from the general population in each country, with older age groups, and higher education typically over-represented^50^. To correct for this representativeness bias, we post-stratified the sample by age, sex, and education level. Each response was weighted to recover the national population characteristics, obtained through the French National Institute of Statistics and Economic Studies (INSEE)^51^ and the Italian National Institute of Statistics (ISTAT)^52^. We considered three age groups (20–44, 45–59, and 60+ years), sex (male and female), and two education attainment levels based on the highest qualification achieved: ‘up to high school’ (including primary, secondary, and high school diplomas) and ‘post-high school’ (including any university studies or vocational training undertaken after high school).

### Statistical analysis

We investigated the determinants of awareness of dengue as a disease, knowledge of its characteristics, adoption of preventive measures, and vaccination willingness. Additionally, we investigated the factors associated with the most common reasons for refusing vaccination if offered: believing oneself to be at low risk of contracting dengue, and not belonging to any risk group. We identified determinants with bias adjusted multivariate logistic and ordinal regression models^53^, depending on the nature of the outcome. In particular, ordinal regression models were used to capture the ordinality of the different levels of knowledge and measure adoption^54,55^. We assessed the assumption of odds proportionality underlying these models in all instances where they were used^56,57^ (see SI). Analyses were conducted on a single pooled dataset including both French and Italian participants. We assessed the homogeneity of the two populations as described in the section *Homogeneity of the two populations.* The analyses were conducted in python using the statsmodels library, together with the R library mass, called in python with r2py.

### Explanatory variables

Explanatory variables, summarised in Table 1, were derived from participants’ intake survey, and from responses to the dengue questionnaire. From the participants’ intake survey, we obtained their country of residence (variable name: *Country*), sex (*Sex*), age (*Age group*), education attainment level (*Education level*), employment status (*Occupation*) and the presence of at least one comorbidity (*Comorbidities*). From the zip code of residence of each participant, we determined the urbanicity of their domicile (*Urbanicity*) using data from ISTAT for Italy^58^ and INSEE for France^59^. We also assigned to each participant a binary variable (*Autochthonous cases*) encoding whether autochthonous transmission of dengue had been reported in their department (France) or province (Italy) of residence during the year 2023^60,61^. Dengue symptom history (*Symptoms*), dengue vaccination status (*Vaccination status*), search of information related to dengue (*Searched information*), knowledge of a case among contacts (*Known case*), level of concern related to dengue among different dimensions (*Worry level*), perceived efficacy of several preventive measures (*Efficacy score*) as well as knowledge of dengue characteristics (*Knowledge score*) and self-evaluated level of information about dengue (*Self-evaluation*) were all obtained from the survey replies.

**Table 1.** Explanatory variables, their levels and reference, as considered in the statistical analysis to explain knowledge, attitudes and practices related to dengue.

| Variable | Values (reference in bold) |
| --- | --- |
| Country | France, Italy |
| Sex | Male, Female |
| Age group | 20-44, 45-64, **65+** |
| Education level | Until high school, **Post high school** |
| Occupation | True, **False** |
| Urbanicity | **Urban**, Rural |
| Symptoms | True, **False** |
| Comorbidities | True, **False** |
| Autochthonous case | True, **False** |
| *Vaccination status* | True, **False** |
| *Searched information* | True, **False** |
| *Known case* | True, **False** |
| *Worry level* | Continuous variable |
| *Knowledge score* | Poor, Medium, Good, **Excellent** |
| *Efficacy score* | Continuous variable |
| *Self-evaluation* | Continuous variable |

The binary variable *Symptoms* was defined to describe participants who had experienced at least two of the following symptoms possibly linked to dengue in the past thirty days: eye pain, blood in the feces, blood in the vomit, bloody gums, and bone pain. We measured *Worry level* as the average of the participants’ concern level across five dimensions, each rated on a 5 points Likert scale from 1 (*Not at all worried*) to 5 (*Extremely worried*). Dimensions included i) fear of being infected, ii) fear of having severe symptoms if sick, iii) fear for friends/family, iv) fear for work/economic consequences in case of contagion, v) fear of work/economic consequences in the event of a pandemic. The *Efficacy score* of a participant was instead computed by averaging their rating of the efficacy of the various measures proposed. Finally, *Self-evaluation* refers to each participant’s personal perceived information level, rated on a scale from 1 (*Very poor or none at all*) to 5 (*Very accurate*).

Participants who were aware of dengue were assigned a *knowledge score* based on the number of known characteristics among the eight presented: Poor (0–2), Medium (3–4), Good (5–6), and Excellent (7–8). The dengue characteristics we enquired about were i) “*Dengue is transmitted through bites of infected A. albopictus (tiger mosquito) or A. aegypti mosquitoes”*, ii) “*Mild cases of dengue have symptoms similar to Covid-19 or the Flu”, iii) “Dengue can be asymptomatic”*, iv) “*Symptoms last on average between 2 and 7 days”*, v) “*Severe cases of dengue can lead to hospitalization and death”*, vi) “*There is no specific treatment or medicine against dengue”*, vii) *“If you have already had dengue in the past, you are more at risk of having severe symptoms”*, viii) “*Pregnant women and children are more at risk of having severe symptoms”*. The variable *knowledge score* was also considered as an outcome variable.

### Outcome variables

#### Awareness of dengue as a disease

Participants were initially asked whether they were “*aware of the existence of an infectious disease called dengue in your country or around the world*”. This outcome was modelled with a bias adjusted logistic regression model, with backward stepwise variable selection (see section *Variable selection*). We included the following variables as predictors prior to variable selection: *Country*, *Sex*, *Age group, Education level*, *Occupation, Urbanicity*, *Symptoms*, *Comorbidities*, and *Autochthonous case*.

#### Knowledge of dengue

We modelled the participants’ *knowledge score* (see section *Explanatory variables)* with a bias adjusted ordinal regression model, with backward stepwise variable selection (see section *Variable selection*). We included the following variables as predictors prior to variable selection: *Country, Sex, Age group, Education level, Occupation, Urbanicity, Symptoms, Comorbidities, Vaccination status, Searched information, Known case, Worry level and Autochthonous case*.

#### Adoption of preventive behaviours

We enquired about 6 specific preventive behaviours known to reduce the risk of contracting dengue^62^: i) “*Wearing long-sleeved clothing covering arms and legs”*, ii) “*Regularly using mosquito repellent”*, iii) “*Avoiding swampy areas or areas with a high density of mosquitoes”*, iv) “*Avoiding traveling/canceling a trip to areas at risk”*, v) “*Installing mosquito nets in all windows”*, vi) “*Removing all stagnant water from the balcony, garden, and windows”*. We considered the total number of adopted measures as an indicator of participants’ prevention practices regarding dengue, binning it in four categories: “poor adoption” (0 measures), “medium adoption” (1-2 measures), “good adoption” (3-4 measures) and “excellent adoption” (5-6 measures).

This outcome was modelled with a bias adjusted ordinal regression model, with backward stepwise variable selection (see section *Variable selection*). We included the following variables as predictors prior to variable selection: *Country, Sex, Age group, Education level, Occupation, Urbanicity, Comorbidities, Vaccination status, Searched information, Known case, Worry level, Knowledge score, Efficacy score and Autochthonous case*.

#### Attitudes towards vaccination

Non-vaccinated participants who had heard of dengue were asked whether they would accept a vaccine if offered. Different reasons for refusal were offered to unwilling participants. We analysed the determinants of vaccination willingness among unvaccinated participants, and the determinants of the two equally most commonly reported reasons for refusing vaccination—“*I don’t think I will get dengue*”, and “*I don’t belong to any risk group*”. These outcomes were modelled with bias adjusted logistic regression models, with backward stepwise variable selection (see section *Variable selection*). We included the following variables as predictors prior to variable selection: *Country, Sex, Age group, Education level, Occupation, Urbanicity, Symptoms, Comorbidities, Searched information, Known case, Worry level, Knowledge score, Self-evaluation and Autochthonous case*.

### Variable selection

For each outcome considered, explanatory variables included in the final models were obtained with a stepwise backward selection process^31,63,64^. Each variable was initially tested in a univariate analysis, and those with a p-value below 0.2 were retained for the multivariate analysis. The least significant variable (highest p-value) was iteratively removed until all remaining predictors had a p-value ≤ 0.05. For categorical variables, the selection was based on the smallest p-value among its levels. For sensitivity, we also considered simultaneously all possible multivariate models for each outcome, and selected the model with highest adjusted pseudo R² (see SI).

### Homogeneity of the two populations

We conducted the statistical analysis on the pooled dataset merging participants from both France and Italy. While we controlled for country of residence by including it as a predictor, the analysis in the absence of interaction term assumes that the effects of the remaining predictors are the same in both countries. For each predictor we tested whether the country of residence had a modulating effect using simple models including interaction terms. For each outcome, we considered a set of logistic or ordinal regression models with the country of residence, the predictor tested for interaction, and their interaction terms. Where the interaction term was significant, we presented the odds ratios separately for the French and Italian cohorts to characterise the effect of the predictor. Where the interaction term was not significant, we concluded that the country of residence had no impact on the effect of the predictor tested, and the two populations were thus homogeneous.

## Results

### Descriptive analysis

#### Participants

Socio-demographic characteristics of the participants considered, before and after bias adjustment, are shown in Table 2. Male individuals were overrepresented in Italy, while the French sample was biased towards females. While more than half French participants were above 65 years old (55%), the most common age group among participants from Italy was “45-64” (51.7% participants). The youngest age-group “20-44” was the least represented in both countries. Participants from Italy were nearly equally likely to have completed pre- or post-high school education. In France instead, the majority of the participants (72%) had educational attainment beyond high school. In order to correct this bias in sex, age and education attainment, we applied post-stratification weights to the samples. Adjusted characteristics of the two samples are shown in Table 2, in the form of percentages. While this process improves the representativeness of the samples, it cannot correct for residual selection bias driven by factors not considered in the stratification.

**Table 2:** Characteristics of the participants sampled. We report raw numbers with their percentage referring to participants in the survey. Percentages in the “Adjusted” columns were obtained with post-stratification adjustment weights.

| Variable | Level | Italy |  | France |  | Total |  |
| --- | --- | --- | --- | --- | --- | --- | --- |
|  |  | Raw | Adjusted | Raw | Adjusted | Raw | Adjusted |
| Sex | Male | 220 (54.5%) | 48.1% | 1060 (42.4%) | 47.5% | 1280 (44.1%) | 47.6% |
|  | Female | 184 (45.5%) | 51.9% | 1440 (57.6%) | 52.5% | 1624 (55.9%) | 52.4% |
| Age group | 20-44 | 78 (19.3%) | 35.1% | 230 (9.2%) | 37.0% | 308 (10.6%) | 36.8% |
|  | 45-64 | 209 (51.7%) | 37.0% | 894 (35.8%) | 35.7% | 1103 (38.0%) | 35.9% |
|  | 65+ | 117 (29.0%) | 27.9% | 1376 (55.0%) | 27.3% | 1493 (51.4%) | 27.4% |
| Education level | Until high school | 196 (48.5%) | 83.8% | 700 (28.0%) | 69.0% | 896 (30.9%) | 71.1% |
|  | Post high school | 208 (51.5%) | 16.2% | 1800 (72.0%) | 31.0% | 2008 (69.1%) | 28.9% |
| Occupation | Yes | 249 (61.6%) | 54.6% | 963 (38.5%) | 54.2% | 1212 (41.7%) | 54.3% |
|  | No | 155 (38.4%) | 45.4% | 1537 (61.5%) | 45.8% | 1692 (58.3%) | 45.7% |
| Urbanicity | Rural | 62 (15.3%) | 15.6% | 677 (27.1%) | 28.2% | 739 (25.4%) | 26.4% |
|  | Urban | 342 (84.7%) | 84.4% | 1823 (72.9%) | 71.8% | 2165 (74.6%) | 73.6% |
| Dengue Awareness | Yes | 356 (88.1%) | 84.2% | 2353 (94.1%) | 92.3% | 2709 (93.3%) | 91.2% |
|  | No | 48 (11.9%) | 15.8% | 147 (5.9%) | 7.7% | 195 (6.7%) | 8.8% |

The remainder of our analysis is conducted on a pooled sample including both sets of participants, unless otherwise specified. All proportions quoted in the rest of this article refer to the adjusted proportions after post-stratification weighting of the samples.

#### Awareness of dengue, and knowledge of its characteristics

The vast majority of respondents (91.2% in the combined sample, after adjustment) had heard of dengue as a disease. This awareness was slightly higher in France (92.3%) than in Italy (84.2%). Knowledge levels however varied widely across different characteristics of dengue (see Figure 1A). While 94.3% of participants recognised that “*dengue is transmitted through the bites of infected A. albopictus (tiger mosquito) or A. aegypti mosquitoes*”, and 88.0% of participants knew that “*Severe cases of dengue can lead to hospitalization and death*”, only 41.0% of respondents were aware that “*dengue can be asymptomatic*”, and just 26.0% knew that “*If you have already had dengue in the past, you are more at risk of developing severe symptoms*”.

A minority (14.8%) of participants had actively looked for information on the spread of dengue in their country, consulting the sources shown in Figure 1B. The most frequently used source was “*Scientific information and dissemination websites*” (69.0% of the participants who had searched for information), followed by “*National newspapers*” (32.8%).

**Figure 1.**
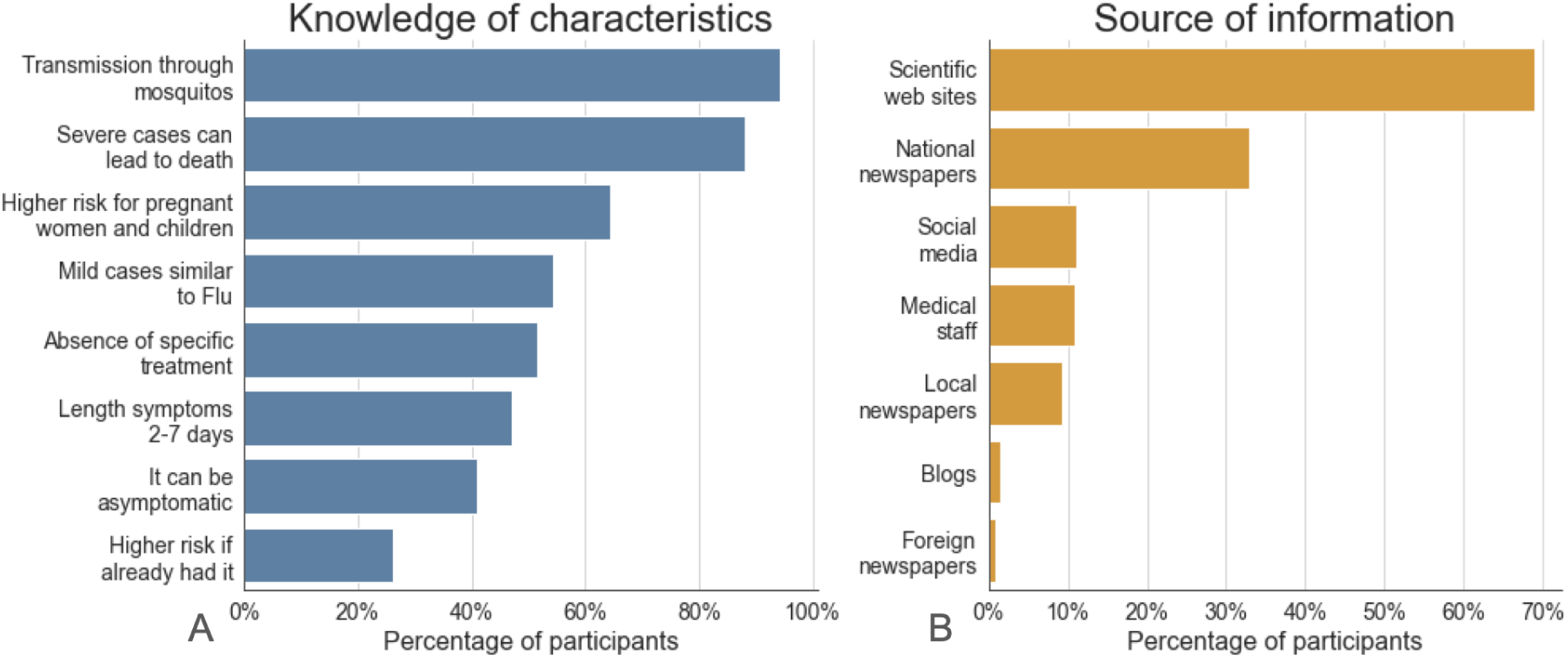
Knowledge among the participants. **A)** Percentage of participants (in the French and Italian samples) knowing each characteristic of dengue **B)** Percentage of participants consulting each source of information. The percentages computed in panel A are with respect to the participants who have heard of dengue (N= 2709). In panel B, the proportions computed are among those participants that have actively sought information on the spread of dengue in their country (N= 419). Percentages are computed using the joint French and Italian samples adjusted with post-stratification weights to recover representative samples in terms of age distribution, sex and education in each country.

#### Adoption and perceived efficacy of preventive measures

The most commonly adopted measure was “*Removing stagnant water from balconies, windows, and gardens*” (44.4%), followed by “*Avoiding swampy areas or areas with a high density of mosquitoes*” (33.2%) (see Figure 2A). In contrast, “*Wearing long-sleeved clothing that covers arms and legs*” and “*Installing mosquito nets on windows*” were adopted by only 16.2% and 15.9% of the participants, respectively. Perceived efficacy of each measure is shown in Figure 2B. All six measures were generally considered effective, with over 80% of respondents rating each measure as at least “quite effective.” For four measures, this percentage exceeded 93%. Interestingly, the two most adopted measures were also perceived as the least effective.

**Figure 2.**
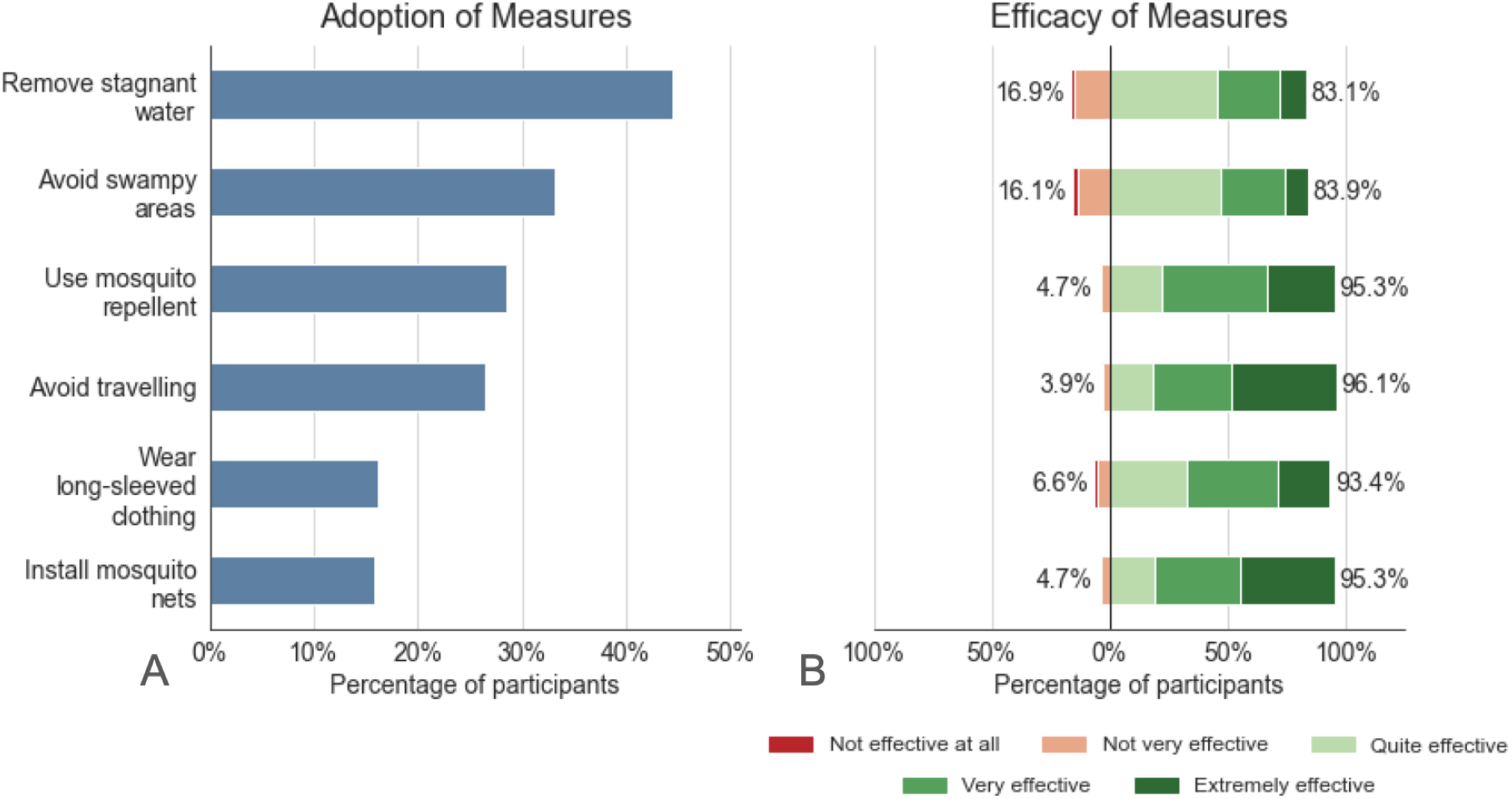
Adoption and perception of preventive measures. **A)** Percentage of participants adopting each preventive measure. **B)** Distributions of perceived efficacy of each measure rated by the participants. In both panels, percentages are computed with respect to the participants who have heard of dengue (N= 2709), using the joint French and Italian samples adjusted with post-stratification weights to recover representative samples in terms of age distribution, sex and education in each country.

#### Fears and vaccination

We observed moderate levels of concerns among the population sampled (see Figure 3A). Concern levels varied slightly depending on consequences of an epidemic. Participants expressed greater concern about severe symptoms and the well-being of friends and family, as indicated by the high percentage of responses in the top three levels of worry. In contrast, work- and economy-related fears —both in the event of infection and in the broader context of an outbreak— showed lower levels of concern. For instance, only 28.2% of participants expressed no concern for “*Fear of having severe symptoms if I get sick*” while 44.3% of participants had no concern for “*Fear of work/economic consequences in case of contagion*.”

Among the 2,709 respondents who had heard of dengue, only 11 had been vaccinated against it. Instead, 59.4% of unvaccinated participants would be willing to receive a vaccine if offered (see Figure 3B). As shown in Figure 3C, the most frequently cited reasons to refuse a vaccine if offered were “*I don’t think I’ll get dengue*”, cited by 37.4% of unwilling participants; and “*I don’t belong to any risk group*”, cited by 37.0%.

**Figure 3.**
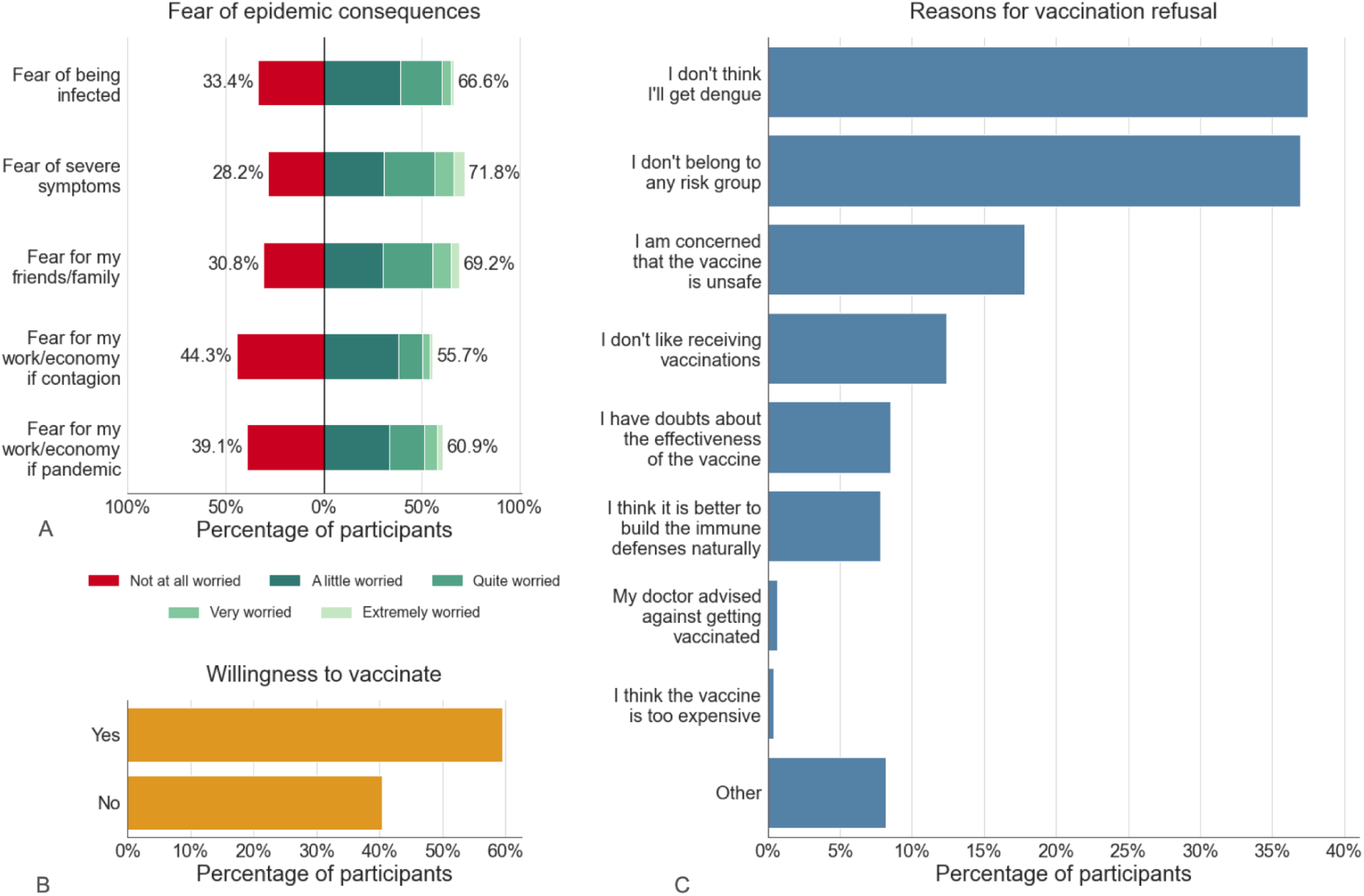
Sentiments towards dengue epidemics, and vaccination against dengue. **A)** Distributions of concern levels for each consequence of a hypothetical dengue epidemic. **B)** Percentage of participants willing to accept a vaccine if offered. **C)** Percentage of unwilling participants citing each reason for refusal. In panel A, percentages are computed with respect to the participants who have heard of dengue (N= 2709). In panel B, percentages are computed with respect to the participants who have heard of dengue and are unvaccinated (N= 2698) while instead percentages reported in panel C are computed with respect to the subset of participants who are unwilling to accept a vaccine (N = 1117). All results shown are obtained using the joint French and Italian samples adjusted with post-stratification weights to recover representative samples in terms of age distribution, sex and education in each country.

### Determinants of knowledge, behaviours and attitudes

Determinants of the awareness of dengue as a disease, knowledge of its characteristics, adoption of preventive measures, and willingness to be vaccinated are shown in Figure 4. We consider odds ratios (ORs) obtained using logistic or ordinal regression models fitted to the joint samples.

**Figure 4.**
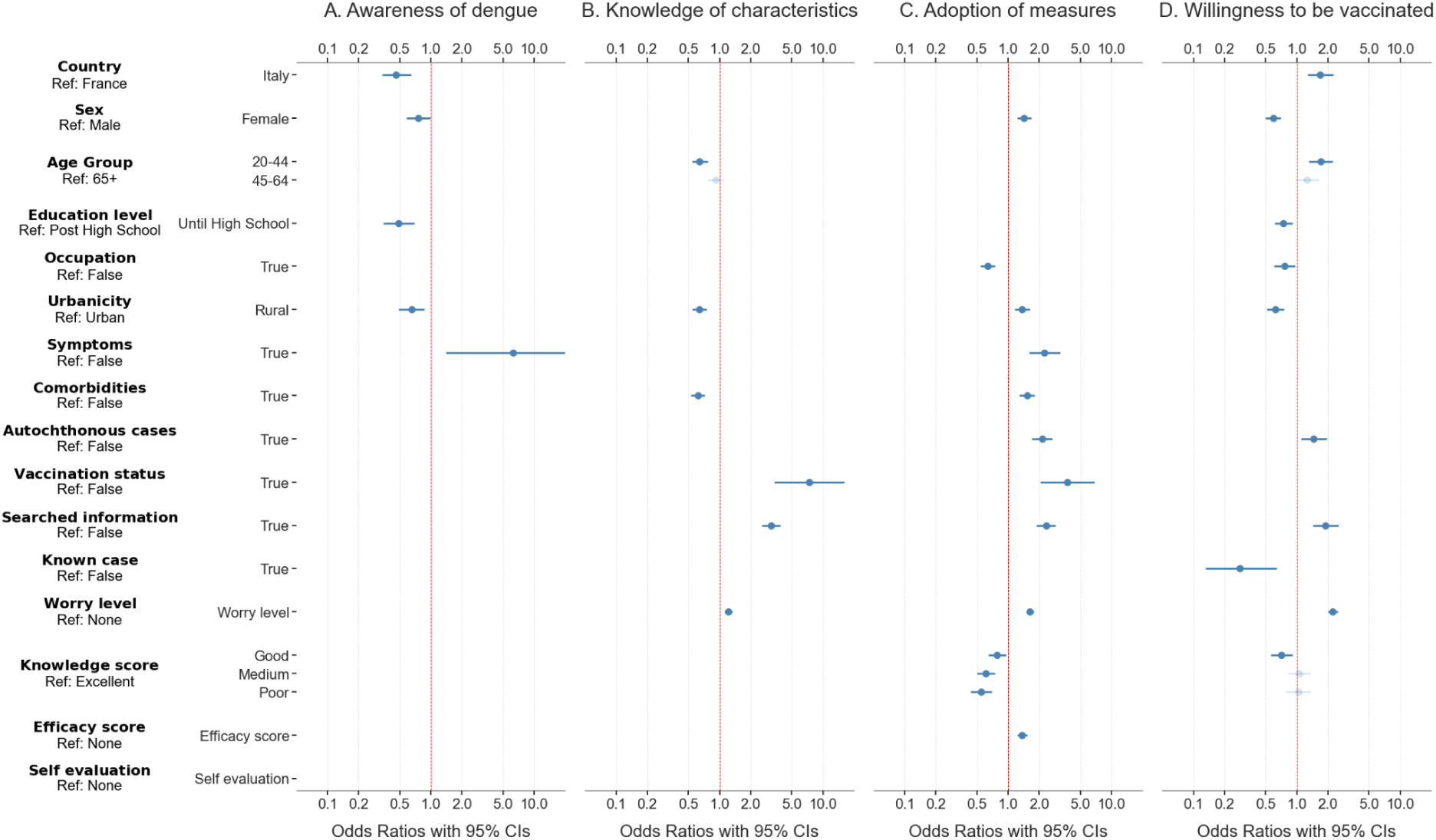
Determinants of knowledge of dengue, its characteristics, adoption of preventive measures, and vaccination willingness. **A)** Determinants of the awareness of dengue identified with a logistic regression model in the form of odds ratios with 95% confidence intervals. **B)** Determinants of the knowledge of dengue characteristics, obtained with an ordinal regression model applied to the answers of the participants who have heard of dengue. **C)** Determinants associated with adoption of preventive numbers (obtained from an ordinal regression model, among participants who have heard of dengue). **D)** Determinants of willingness to be vaccinated (binary logistic model) among the participants who are unvaccinated. Models are multivariate, with variables selected as described in the Methods section. Variables without an associated OR for a given outcome were either not included as explanatory variables in the initial list of considered determinants, or were excluded during the variable selection process. Faded points represent non significant levels of included variables. All analyses are conducted on the pooled dataset including French and Italian populations.

### Awareness of dengue

Our logistic regression model for awareness of “*the existence of an infectious disease called dengue*” identified sex, country of residence, urbanicity, education attainment, and symptom history as drivers for awareness (see Figure 4A). Being female (OR 0.76), living in Italy (OR 0.46), living in a rural area (OR 0.65) and low education attainment level (OR 0.49) were all characteristics associated with lower awareness. Instead, participants who had experienced at least two of the five considered symptoms possibly linked to dengue in the past thirty days were more likely to have heard of the disease (OR 6.29). We found no associations between awareness of the disease and presence of comorbidities, occupation, age group, or living in a department/province with reported locally acquired dengue cases in the past year.

### Knowledge of characteristics

Drivers associated with better knowledge of dengue characteristics, among the participants who had heard of dengue as a disease (N = 2709), are shown in Figure 4B. Knowledge appeared associated in particular with information seeking behaviour (OR 3.17), concern levels (OR 1.21), and previous vaccination against dengue (OR 7.4). Urbanicity also played a role, with participants in rural areas having lower odds of high knowledge levels (OR 0.64) with respect to urban residents. Knowledge instead was not significantly determined by participants’ country of residence, sex, education attainment, occupation, symptom history, knowledge of a dengue case or living in an area with autochthonous transmission in 2023.

### Adoption of measures

Adoption of preventive measures was associated with a combination of behavioural and socio-demographic determinants (see Figure 4C). Perceived efficacy of the measures (OR 1.36), concern levels (OR 1.63), and having actively sought information (OR 2.33) were all associated with higher odds of adopting preventive measures. Individuals with “good”, “medium” and “poor” knowledge all had lower odds of adopting preventive measures (OR = 0.78, OR = 0.61, and OR = 0.55, respectively) compared to individuals with “excellent” knowledge. The local context was also associated with the adoption of preventive measures. In particular, residing in a rural area (OR = 1.36) and living in a province/department with previous local transmission of dengue (OR = 2.13) were both associated with higher odds of adopting preventive measures. Female participants had 1.43 times the odds of adopting a larger number of measures than men. Previous vaccination and dengue symptom history were also positively associated with higher adoption of measures. Being employed had the opposite effect, and was associated with lower odds of adopting a larger number of measures (OR = 0.63). Adoption of measures however did not significantly depend on country of residence, education attainment, and knowledge of a case of dengue among contacts.

### Willingness to vaccinate

Focusing on unvaccinated participants (N = 2698), we identified determinants associated with willingness to accept a vaccine if offered (see Figure 4D). Socio-demographic and contextual characteristics were especially associated with willingness. Higher willingness was associated with residing in Italy (OR 1.69), the 20–44 age group (OR 1.70), and areas where locally acquired cases of dengue were reported in 2023 (OR 1.46). Instead, female sex (OR 0.59), education attainment limited to high-school (OR 0.74), having an occupation (OR 0.76), and living in rural areas (OR 0.62) was associated with lower willingness. Behaviours and attitudes were also correlated to willingness: information seeking (OR 1.90), and increased concerns (OR 2.23) were associated with higher willingness. Willingness was independent of participants’ own perceived level of information, comorbidities and symptom history.

### Reasons for vaccination refusal

We identified determinants explaining the most commonly cited reasons for vaccine refusal among unwilling participants (N= 1117) (see Figure 5). Both *“I don’t think I will get dengue”* and *“I don’t belong to any risk groups”* were less likely to be cited by participants with higher concerns (OR 0.65 and 0.76 respectively). Residing in Italy (OR 1.64), having an occupation (OR 1.54), and having a “good” (compared to “excellent”) knowledge of dengue characteristics (OR 1.50) showed higher odds of refusal with “*I don’t belong to any risk group*”. Instead, the 20-44 age group (OR 0.47), education attainment limited to high-school (OR 0.53), and comorbidities (OR 0.32) were associated with lower odds of citing this reason. Italian residents (OR 0.34), those with an occupation (OR 0.61), and those perceiving themselves more highly informed (OR 0.73) had lower odds to quote *“I don’t think I’ll get dengue*”, while residing in urban areas instead increased the odds of citing this reason (OR 1.36). The remaining of the variables represented in Figure 5 without an associated OR were not found to be significant by our analysis.

**Figure 5.**
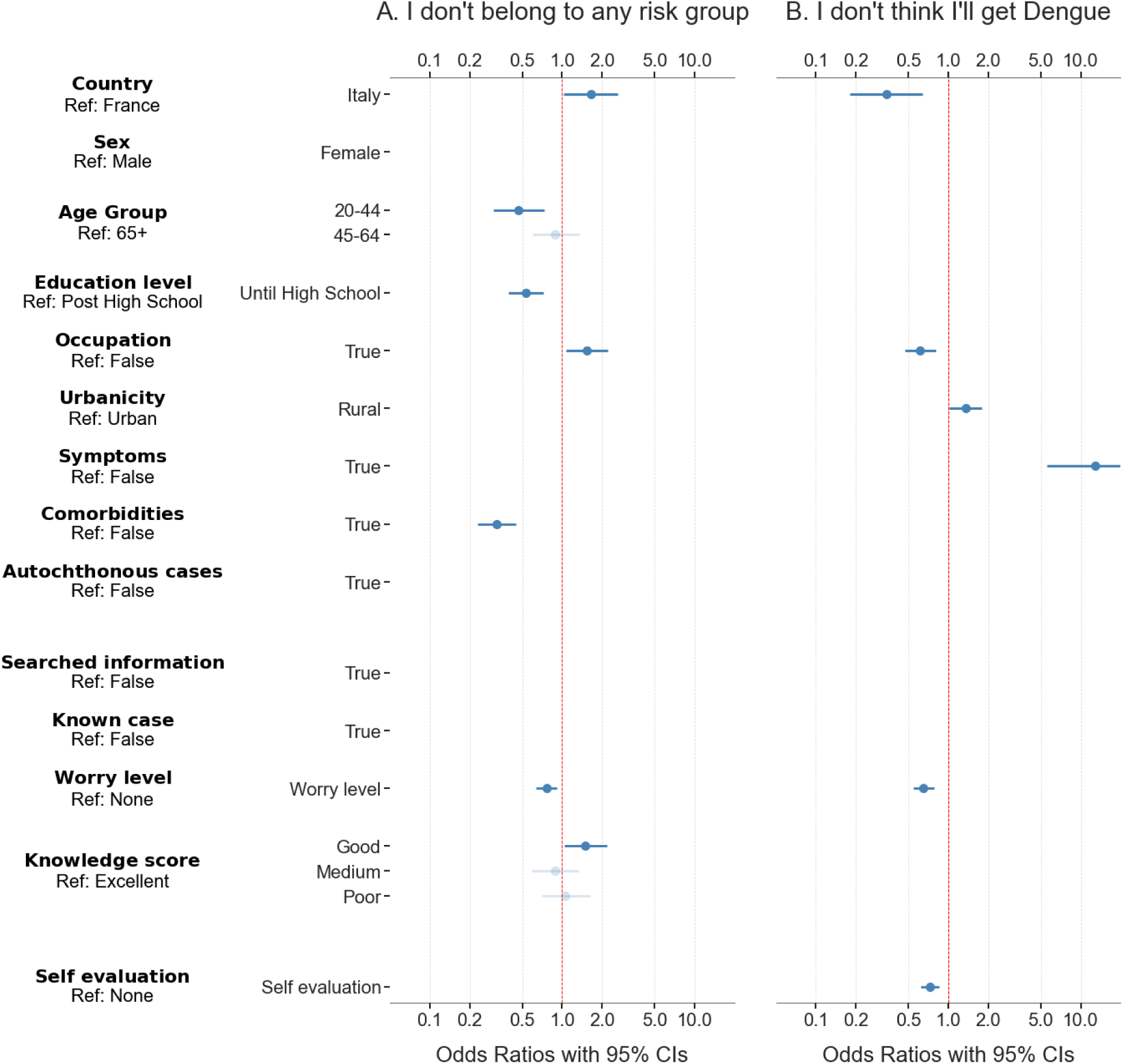
Drivers of the main reasons for vaccine refusal. **A)** Determinants of citing “I don’t belong to any risk group” in the form of odds ratios with 95% confidence intervals. **B)** Determinants of citing “I don’t think I’ll get dengue” in the form of odds ratios with 95% confidence intervals. Odds ratios were obtained from a multivariate, bias adjusted logistic regression model applied to the participants who are unwilling to be vaccinated. Variables without an associated OR were excluded during the variable selection process. Faded points represent non significant levels of included variables. All analyses are conducted on the pooled dataset including French and Italian populations.

### Interactions between the country of residence and the identified determinants

We refer to the Supplementary Information for results of the bivariate regression analyses assessing the homogeneity of the French and Italian samples. In the majority of the determinants considered, interaction terms with the country of residence were not significant, indicating that the determinant’s effects apply independently of countries. Where interaction terms were statistically significant, the main effect of country of residence (i.e., the Italian dummy variable) often became non-significant. This may be due to the fact that the independent effect of the country was absorbed into the specific interaction terms, indicating that the effect of those predictors was not independent of country but rather country-specific.

Interactions were identified in a few cases, highlighting few cross country differences (see SI). Among Italian participants, those with an occupation were less likely to know a higher number of characteristics than those without, whereas in France employment status did not affect knowledge. While the main analysis shows that urbanicity was negatively associated with adopted measures, this reversed in Italy. A similar effect was measured in the case of the association between areas where autochthonous cases had been reported, and willingness to be vaccinated. The first effect may reflect structural differences in the rural–urban context between the two countries, while the second could be due to the small number of distinct areas with autochthonous cases in Italy in 2023. Finally, while sex was not associated with “*I don’t think I’ll get dengue*” in the joint analysis, female participants were less likely than males to report this reason in France, but instead more likely in Italy.

### Variance Inflation Factor

As detailed in the Supplementary Materials, all explanatory variables included in the final models were tested for multicollinearity using the Variance Inflation Factor (VIF). All VIF values were below 3, with the exception of three variables (3 < values < 10), indicating that multicollinearity was not a concern in our analyses.

## Discussion

Using French and Italian participatory surveillance cohorts of the European network InfluenzaNet^45^, we gathered data on knowledge, attitudes and preventive practices related to dengue in two countries where dengue is an emerging threat. Interestingly, we found high awareness, moderate levels of concerns, and a moderate vaccination willingness. Knowledge and attitudes, socio-economic factors and the context explained the adoption of preventive measures, suggesting targeted approaches to increase adoption. Moreover, this study shows how pre-existing participatory surveillance cohorts can be pivoted towards emerging threats to assess knowledge, awareness and attitudes, as well as to evaluate determinants of knowledge, behaviours, and attitudes towards vaccination in order to help design efficient interventions.

Several surveys assessed dengue awareness and knowledge in non-endemic regions since the first detections of autochthonous cases^33,35^. In mainland France, successive survey waves probed the population between 2012 and 2014, finding increased knowledge that dengue can be transmitted by mosquitoes (from 58.5% in 2012 to 76.2% in 2014), but stable perceptions of severity and vulnerability^38,39^. Evidence from the French public health agency (Santé Publique France) however found that only 28.2% of the population surveyed in metropolitan areas with *A. albopictus* presence knew that mosquitoes around them could transmit dengue^37^, suggesting that knowledge may not translate to risk and susceptibility awareness. In the Lazio region of Italy, while 49.8% of Italian participants knew that tiger mosquitoes could transmit “some diseases”, only 1% was able to correctly identify which ones in 2016. This reached 28.7% among Lazio residents originally from Kerala (South India) where dengue is endemic, showing wide gaps in knowledge between communities, and the crucial impact of the social and cultural context^36^. Similarly, we noted an association between vaccination status and knowledge in our analysis. As vaccination is not currently recommended to the general population, vaccinated individuals included in the French sample might have received the vaccine while residing in overseas departments where dengue is endemic, which may have shaped their knowledge. Finally, our analysis conducted on a sample collected in 2024 in Italy and France suggests higher awareness levels about the existence of dengue (91.2%) and higher knowledge of its transmission mode (94.3%), possibly due to increasing reports of local transmissions since previous surveys were implemented.

Our survey focused on the knowledge of characteristics specific to dengue, while existing research was typically centred on knowledge related to mosquitoes and the diseases they carry^35,36,38–40^. Heterogeneous knowledge of symptoms of mosquito-borne viruses has been previously noted in France in 2012^39^. Our study found persistent heterogeneity in knowledge of dengue characteristics in 2024: the transmission mode of dengue was known by 94.3%, while only 26.0% of participants were aware of the increased risk associated with reinfection, and 41.0% knew that dengue could be asymptomatic.This shows that, while some features of dengue are highly known to populations even in non endemic regions, some clinical signs of the disease and risk factors remain poorly assessed by the population. This highlights an urgent need to implement communication campaigns to raise knowledge and awareness of these lesser known characteristics.

In particular, efforts should focus on the younger population, rural communities, and less educated groups, which were less likely to have heard of the disease, or know its characteristics. Moreover, the lower knowledge of characteristics among those with comorbidities suggests that information campaigns in medical establishments, often attended by this subpopulation, could be beneficial. Increasing knowledge in the population is particularly important, as higher knowledge is known to be associated with the adoption of preventive measures^31,34,40^. This is also the case in our sample, confirming that information campaigns raising knowledge could increase preventive measure adoption, and thus lead to better vector control. Understanding information source preferences can suggest specific communication channels that can be used to raise awareness, improve knowledge and encourage the adoption of preventive practices. In Spain and the Netherlands, 93.7% and 81.6% of participants were interested in receiving information on mosquitoes and mosquito-borne viruses^40^. Preferred sources however varied between countries, indicating that the local context must be taken into account to best reach the population^40^. Our survey showed that 14.6% of individuals had searched for information on the circulation of dengue, most often from scientific websites, followed by national newspapers. Future campaigns should therefore leverage these channels of communication to inform the population, but also invest in alternatives to better reach those individuals that do not spontaneously search for information. The identification of these alternatives should be the focus of future research.

Our findings suggest a higher trend in the adoption of preventive practices compared to previous survey results from the past decade. After the first detection of locally transmitted dengue cases in Europe in 2012, approximately 50% of participants surveyed in France were adopting at least one recommended preventive measures^39^. Similarly, 47% of Italians surveyed in 2016 reported adopting precautions against tiger mosquitoes^36^. More than a decade later, our survey shows that the adoption of at least one preventive measure in France and Italy has increased to 59%. This increasing trend can be especially observed for eliminating standing water which was adopted by 17.7% of participants in 2012^39^, and 44.4% in our study, and repellent use which was adopted by 17.4% of participants in 2012^39^, 20.3% in 2016^37^ and 28.5% in our study in 2024. Use of mosquito nets and long sleeved clothing however remained stable since 2016, with 17.9% of participants reporting using nets often in 2016^37^, close to the 15.9% of the participants in our study. Similarly, long sleeved clothing was used often by 14.1% of the participants in 2016^37^ compared to 16.2% in our study. The most adopted measure was also considered to be the least efficient by the participants in our study. This disconnect between perceived efficacy and adoption holds for few of the measures we considered, and was also reported in La Réunion where dengue is endemic^30^. This could stem from the varying efforts required to adopt each measure, and the fact that the adoption of each measure may depend on participants’ habitat and lifestyle habits.

In line with expectations from the Health Belief Model^28^, the adoption of preventive measures in our sample was the result of a complex interplay between knowledge of dengue and attitudes, beliefs, socio-economic and contextual factors. In particular, raised concern levels driven in part by perceived susceptibility and severity, perceived efficacy of measures (akin to perceived benefits), as well as several individual characteristics (being female, not having an occupation, having comorbidities, being vaccinated and living in a rural area) increased the likelihood of adopting a larger number of preventive measures in our study. Moreover, previous symptom history and the detection of local cases appeared as cues to action which increased the likelihood of measures adoption, together with having sought information. These results confirm previous findings highlighting the role of sex, knowledge, concern levels, perceived efficacy and education status in shaping the adoption of preventive measures against dengue and other mosquito-borne diseases^37–40^. In addition, our study also revealed the role of the participants’ information search behaviour, occupation and urbanicity on the adoption of preventive measures.

Furthermore, the drivers of adoption of preventive measures identified by our study are partially in line with known drivers of preventive behaviours against COVID-19^65^, such as masking, self isolating when symptomatic, social distancing or increased hygiene. The effect of sex especially has been extensively investigated, with women repeatedly found more likely to engage in preventive behaviours during the COVID-19 pandemic^65–67^. Similarly, perceived efficacy of measures, and unemployment are both predictors of adoption of preventive behaviour against dengue in our study, and during the COVID-19 pandemic^66,68,69^. However, older age and higher education levels, which were often associated with the adoption of measures during the COVID-19 pandemic^65,67,69^, were not significant factors in the context of dengue in our study. On the other hand, the higher likelihood of adopting preventive behaviours against COVID-19 in urban settings^70^ is reversed in the context of dengue. This reversal could stem from the fundamental difference in preventive measures between the two diseases: common preventive measures against COVID-19 such as wearing a mask in public transport, working from home or avoiding restaurants may be more applicable in urban settings while preventive measures against dengue such as removing stagnant water or using mosquito nets may be more applicable instead in rural settings.

Nonetheless, the lower likelihood of adopting preventive measures against dengue in urban settings is a particularly concerning finding, as *A. aegypti* and *albopictus* mosquitoes adapted to thrive in urban environments where they face fewer predators, have access to a high density of human hosts, and many man-made structures where they can breed^71,72^. Future campaigns to encourage the uptake of preventive measures in France and Italy should therefore aim to raise knowledge, efficacy perceptions, inform on dengue severity and susceptibility, and should focus in particular on informing urban residents on the actions they can take to protect themselves against dengue.

Our survey suggests that concern levels related to dengue fever remain moderate in France and Italy, despite the increasing number of cases reported^21^. Our study further sheds light on the main causes of concerns for individuals sampled. Fears of developing severe symptoms was the leading cause of worries in participants, followed by fears for close ones and relatives and fear of being infected. Participants were less worried about economic consequences due to contagion or a dengue epidemic. In line with findings from several empirical studies, this shows that dengue and more generally mosquito-borne diseases are not perceived as a significant threat in European populations^36,37,39,41^, including by health professionals as evidenced by the low risk projections of specialised French medical doctors^42^. This is at odds with the anticipated increased risk of local outbreaks^25^ due to the expansion of the vector on the European continent^18,19^, and increase in climatic suitability for local spread^20^.

Our study investigated attitudes towards vaccination against dengue in a non-endemic context. In line with the restricted eligibility for vaccination in Italy and mainland France, we observed a very low vaccination coverage in our sample, with only 11 vaccinated participants in total. Our survey evidenced a high vaccination willingness in the hypothetical scenario of a vaccine being offered, especially in light of the moderate concerns expressed about the consequences of dengue outbreaks. The willingness in the general French and Italian populations may be different from our estimates coming from participatory cohorts, who typically demonstrate higher vaccination coverage than the general population^50^. However, our analysis suggests that future vaccines against dengue would be well received in France and Italy.

Willingness to be vaccinated was however higher in Italy than France, as observed previously before the COVID-19 vaccine roll-out^73^. We also identified a significant sex divide, with women less likely to accept vaccination, in line with numerous evidence on the impact of sex on vaccination willingness to prevent respiratory viruses^74,75^. Interestingly, we found that younger individuals were more likely to accept a vaccine, in contrast with findings related to respiratory viruses with a strong age dependent severity profile^74^. While dengue is also more severe in the elderly^76^, this does not translate into higher vaccination willingness in this group. Higher education attainment and employment were also associated with higher vaccination willingness. The local environment (urbanicity, and the detection of locally transmitted cases previously) also positively influenced the hypothetical uptake together with concern levels. Our findings on the negative impact of the knowledge of a case on vaccination willingness is unexpected. This effect deserves further investigation. Instead, reasons to refuse vaccination most often cited were related to risk perception, either through perceived susceptibility (*“I don’t think I will get dengue”*) or perceived severity (*“I don’t belong to any risk group”*). Our analysis suggested the importance of the broader social context in shaping vaccination refusal attitudes through the country of residence. In both cases, individuals who reported higher concerns for dengue consequences in the survey were less likely to cite either reason. Little overlap in determinants were found, suggesting the unique interplay of individual characteristics shaping each refusal reason.

Novel insights, such as those presented here, can be obtained rapidly and at low cost by polling volunteers already engaged in participatory surveillance cohorts. The platforms in the InfluenzaNet network were initially designed for influenza-like illnesses surveillance^45^, and rapidly evolved during the pandemic to capture COVID-19 illnesses. This was instrumental to evaluate knowledge, attitudes and practices at the beginning of the first wave of COVID-19 in March 2020 in several European countries^49^, and later on, to reassess attitudes towards testing and boosting vaccination as the context evolved^46^. With the present study, we illustrate how participatory surveillance cohorts can also be leveraged to investigate emerging threats, unrelated to respiratory viruses. In the future, these platforms could be used to investigate the evolution of knowledge, attitudes and practices regarding dengue, as well as other vector borne diseases. These findings could also be analysed through additional indicators related for example to information exposure^77^. Finally, future efforts remain necessary to understand the drivers of exposure to dengue, related to individual habitats, lifestyle and behaviours, and their link to knowledge, attitudes, and the adoption of preventive measures.

Our study has several limitations. First, participatory surveillance exhibits an intrinsic selection bias in the samples. As participation is voluntary, the population surveyed may be more likely to engage with health recommendations, protective behaviours, and to consider vaccination positively. This bias could only be corrected by using different costly collection methods such as representative panels or door to door collection. Moreover, participants to InfluenzaNet cohorts participate online, limiting the representation of individuals with limited access to the internet in the sample. Still, all age classes, sex, and education levels were represented in our sample. Despite post-stratifying by age, sex, and education attainment, the representativeness of health related attitudes and behavior may be limited. For example, influenza vaccination coverage in InfluenzaNet cohorts is slightly higher than in the general population^78,79^ while incidence trends of influenza-like illnesses obtained from InfluenzaNet cohorts are consistent with ECDC trends in terms of timing and magnitude^78^. Individual risk factors for influenza-like illnesses obtained from InfluenzaNet cohorts have also been demonstrated to be consistent with the literature^78^. Second, all preventive measures were considered for all participants, regardless of their relevance in each participant’s individual context. For example, individuals in apartments may not have any opportunity to remove stagnant water or avoid swampy areas, while participants who rarely travel may not need to avoid travelling to at-risk areas. In order to mitigate this, we considered the total number of adopted measures as on outcome, instead of each measure individually. Although the questions specifically addressed measures to prevent dengue infection, we also cannot rule out that some participants reported actions motivated instead by a desire to reduce mosquito nuisance^40^. Our questionnaire did not address participants’ perceptions towards mosquitoes (discomfort felt with bites, perceived biting frequency, perceived *Aedes* mosquito presence), which were shown in 2012 to be associated with the adoption of preventive behaviours^37,39^. This factor, among other unaccounted for characteristics, such as type of habitat, household composition, and cultural background from dengue-endemic regions, may act as confounders in our analysis. Future data collection using longer and more detailed surveys may address these aspects.

## Conclusions

Participatory surveillance cohorts provide a unique opportunity to probe engaged participants on their knowledge, attitudes, behaviours and practices related to dengue. We surveyed 2,904 participants of the French and Italian cohorts of the InfluenzaNet network in 2024. Our analysis demonstrates a high awareness of the existence of dengue, but a heterogeneous knowledge of its different characteristics, suggesting an urgent need to raise awareness and educate populations on lesser known aspects. Statistical determinants of awareness, knowledge, adoption of preventive measures and attitudes towards vaccination (willingness, and reasons for refusal) revealed a rich interplay between knowledge levels, concerns, attitudes, behaviours, as well as socio-demographic and contextual characteristics. Notably, active information search and higher concern levels were found to correlate with knowledge, adoption of measures, and willingness to vaccinate. Knowledge in turn was associated with adoption of preventive measures, alongside perceived efficacy, local epidemiological context, occupation of participants, sex, vaccination and symptom history. Targeted communication strategies accounting for these factors are essential to enhance preparedness and outbreak prevention in these regions.

## Data Availability

The raw data supporting the findings of this paper cannot be shared publicly as it contains personal individual information which could compromise users privacy. Data requests should be addressed to the corresponding author.

## List of abbreviations

A: albopictus – Aedes albopictus
A: aegypti – Aedes aegypti
SI: Supplementary Information COVID-19 – Coronavirus disease 2019
ECDC: European Center for Disease Control ISTAT – Italian National Institute of Statistics
INSEE: French National Institute of Statistics and Economic Studies OR – Odds ratio
VIF: Variance Inflation Factor CI – Confidence Interval

## Declarations

### Ethics approval and consent to participate

This study was conducted in agreement with country-specific regulations on privacy and data collection and treatment. Informed consent was obtained from all participants enabling the collection, storage, and treatment of data, and their publication in anonymized, processed, and aggregated forms for scientific purposes. In addition, approvals by Ethical Review Boards or Committees were obtained, where needed according to country-specific regulations.

In France, the Grippenet/Covidnet study was approved by the Comité consultatif sur le traitement de l’information en matière de recherche (CCTIRS, Advisory committee on information processing for research, authorization 11.565) and by the Commission Nationale de l’Informatique et des Libertés (CNIL, French Data Protection Authority, authorization DR-2012-024).

For influweb.it the research was conducted in accordance with Italian Data Protection Authority (Garante per la protezione dei dati personali) regulations on privacy, data collection and treatment, reported here https://www.garanteprivacy.it/web/garante-privacy-en. The Italian Data Protection Authority adheres to the norms set by the European Union’s General Data Protection Regulation (GDPR). Influweb’s data collection process was reviewed and approved by the institutional review board of the ISI Foundation which waived the need for ethics approval as the following applies: all use takes place in compliance with the rules contained in the GDPR; informed consent was obtained online from all participants of the platform at enrollment according to regulations, enabling the collection, storage, and treatment of data, and their publication in anonymized, processed, and aggregated forms for scientific purposes; the website has a “Privacy Statement” section in which the users who decide to enroll in the study can find all the information on who is responsible for the data acquisition and processing in each country. Informed consent was obtained online from all participants enabling the collection, storage, and treatment of data, and their publication in anonymized, processed, and aggregated for scientific purposes. The Influweb website (https://influweb.org/) has a “Privacy Policy” section in which the users who decide to enroll in the study can find all the information on who is responsible for the data acquisition and processing. To ensure privacy and data security, all collected data is pseudonymized—participants’ personal identifiers (e.g., email addresses) are stored separately and are inaccessible to researchers. Survey responses are linked only via an anonymized ID, preserving full privacy. Influweb operates on Google Cloud Platform (GCP) infrastructure in Europe, maintaining GDPR compliance, with continuous backups. Additionally, security measures to protect participant data include hashed passwords, two-factor authentication (2FA) and protection of data in transit by Transport Layer Security v1.3 (“TLSv1.3”).

## Availability of data and materials

The raw data supporting the findings of this paper cannot be shared publicly as it contains personal individual information which could compromise users’ privacy. Data requests should be addressed to the corresponding author.

## Authors’ contributions

MM, NG and DP conceived the study. MM, NG, DP, AB, NP, ADG designed the original survey. Data were processed and prepared for analyses by CT and RM. All authors contributed to the design of the study protocol. ADG and LC performed the statistical analysis, and produced the first draft of the manuscript. All authors contributed to the interpretation of the results, and reviewed, edited, and approved the final version.

## Acknowledgments

The authors thank the “Workshop on Scientific Evolutionary Writing” held in Rome in September 2024, during which the Introduction was drafted. The authors are grateful to Camille Coustaury, Caroline Guerrisi, Anthony Hauser and Olivier Steichen for useful discussions and suggestions on the statistical analyses conducted. The authors are grateful to Chiara Sabbatini for helping translate the Italian survey to French, and Audrey Le Hegaret for helping to implement the survey on Grippenet/Covidnet.

## References

1. Paz-Bailey, G., Adams, L. E., Deen, J., Anderson, K. B. & Katzelnick, L. C. Dengue. The Lancet 403, 667–682 (2024).

2. Bhatt, S. et al. The global distribution and burden of dengue. Nature 496, 504–507 (2013).

3. James, S. L. et al. Global, regional, and national incidence, prevalence, and years lived with disability for 354 diseases and injuries for 195 countries and territories, 1990–2017: a systematic analysis for the Global Burden of Disease Study 2017. The Lancet 392, 1789–1858 (2018).

4. Roth, G. A. et al. Global, regional, and national age-sex-specific mortality for 282 causes of death in 195 countries and territories, 1980–2017: a systematic analysis for the Global Burden of Disease Study 2017. The Lancet 392, 1736–1788 (2018).

5. Scott, L. J. Tetravalent Dengue Vaccine: A Review in the Prevention of Dengue Disease. Drugs 76, 1301–1312 (2016).

6. Vaccines and immunization: Dengue. https://www.who.int/news-room/questions-and-answers/item/dengue-vaccines. [online, accessed: 05/11/2025]

7. Casagrande Passoni Lopes, L., et al. Dengue prevention and treatment: a scoping review. Rev Patol Trop 53, 251–266 (2024).

8. World Health Organization. Dengue guidelines for diagnosis, treatment, prevention and control : new edition. https://iris.who.int/handle/10665/44188 (2009). [online, accessed: 05/11/2025]

9. Pang, T., Mak, T. K. & Gubler, D. J. Prevention and control of dengue—the light at the end of the tunnel. The Lancet Infectious Diseases 17, e79–e87 (2017).

10. Rather, I. A. et al. Prevention and Control Strategies to Counter Dengue Virus Infection. Front. Cell. Infect. Microbiol. 7, (2017).

11. Dambach, P., Louis Valérie R., Standley, Claire J. & and Montenegro-Quiñonez, C. A. Beyond top-down: community co-creation approaches for sustainable dengue vector control. Global Health Action 17, 2426348 (2024).

12. Andersson, N. et al. Evidence based community mobilization for dengue prevention in Nicaragua and Mexico (Camino Verde, the Green Way): cluster randomized controlled trial. BMJ h3267 (2015) doi:10.1136/bmj.h3267.

13. Fotakis, E. A. et al. Description and comparison of national surveillance systems and response measures for Aedes-borne diseases in France, Italy and Portugal: a benchmarking study, 2023. Eurosurveillance 30, 2400515 (2025).

14. Brady, O. J. et al. Refining the Global Spatial Limits of Dengue Virus Transmission by Evidence-Based Consensus. PLOS Neglected Tropical Diseases 6, e1760 (2012).

15. Rocklöv, J. & Dubrow, R. Climate change: an enduring challenge for vector-borne disease prevention and control. Nat Immunol 21, 479–483 (2020).

16. Islam, J., Frentiu, F. D., Devine, G. J., Bambrick, H. & Hu, W. A State-of-the-Science Review of Long-Term Predictions of Climate Change Impacts on Dengue Transmission Risk. Environ Health Perspect 133, 056002 (2025).

17. Ebi, K. L. & Nealon, J. Dengue in a changing climate. Environmental Research 151, 115–123 (2016).

18. Fischer, D., Thomas, S. M., Niemitz, F., Reineking, B. & Beierkuhnlein, C. Projection of climatic suitability for *Aedes albopictus* Skuse (Culicidae) in Europe under climate change conditions. Global and Planetary Change 78, 54–64 (2011).

19. Liu-Helmersson, J., Rocklöv, J., Sewe, M. & Brännström, Å. Climate change may enable *Aedes aegypti* infestation in major European cities by 2100. Environmental Research 172, 693–699 (2019).

20. Liu-Helmersson, J. et al. Climate Change and Aedes Vectors: 21st Century Projections for Dengue Transmission in Europe. eBioMedicine 7, 267–277 (2016).

21. Naddaf, M. Dengue is spreading in Europe: how worried should we be? Nature 10.1038/d41586-023-03407-6 (2023) doi:10.1038/d41586-023-03407-6.

22. Franke, F. et al. Autochthonous chikungunya and dengue fever outbreak in Mainland France, 2010-2018. European Journal of Public Health 29, ckz186.628 (2019).

23. Sacco, C. et al. Autochthonous dengue outbreak in Marche Region, Central Italy, August to October 2024. Eurosurveillance 29, 2400713 (2024).

24. Chikungunya, dengue et zika - Données de la surveillance renforcée en France hexagonale 2024. https://www.santepubliquefrance.fr/maladies-et-traumatismes/maladies-a-transmission-vectorielle/chikungunya/articles/donnees-en-france-metropolitaine/chikungunya-dengue-et-zika-donnees-de-la-surveillance-renforcee-en-france-hexagonale-2024. [online, accessed: 05/11/2025]

25. European Centre for Disease Prevention and Control. Public Health Guidance for Assessing and Mitigating the Risk of Locally-Acquired Aedes-Borne Viral Diseases in the EU/EEA. (Publications Office, LU, 2025).

26. Recrudescence de cas importés de dengue en France hexagonale : appel à la vigilance à l’approche de la saison d’activité du moustique tigre. https://www.santepubliquefrance.fr/les-actualites/2024/recrudescence-de-cas-importes-de-dengue-en-france-hexagonale-appel-a-la-vigilance-a-l-approche-de-la-saison-d-activite-du-moustique-tigre. [online, accessed: 05/11/2025]

27. Rapport au nom de la commission d’enquête chargée d’évaluer les recherches, la prévention et les politiques publiques à mener contre la propagation des moustiques Aedes et des maladies vectorielles. (2020). https://www.assemblee-nationale.fr/dyn/15/organes/cnpe/commission-d-enquete-sur-la-lutte-contre-la-propagation-des-moustiques-aedes [online, accessed: 05/11/2025]

28. Janz, N. K. & Becker, M. H. The Health Belief Model: A Decade Later. Health Education Quarterly 11, 1–47 (1984).

29. Vu, S. N. et al. Elimination of dengue by community programs using Mesocyclops(Copepoda) against Aedes aegypti in central Vietnam. Am J Trop Med Hyg 72, 67–73 (2005).

30. Lamaurt, F. et al. Knowledge, Attitudes, Beliefs, and Practices Regarding Dengue in La Réunion Island, France. International Journal of Environmental Research and Public Health 19, 4390 (2022).

31. Elsinga, J. et al. Knowledge, Attitudes, and Preventive Practices Regarding Dengue in Maracay, Venezuela. Am J Trop Med Hyg 99, 195–203 (2018).

32. Chan, E. Y. Y. et al. Sociodemographic predictors of knowledge, mosquito bite patterns and protective behaviors concerning vector borne disease: The case of dengue fever in Chinese subtropical city, Hong Kong. PLOS Neglected Tropical Diseases 15, e0008993 (2021).

33. Jahromi, A. S. et al. Global systematic review and meta-analysis of knowledge, attitudes, and practices towards dengue fever among the general population. Asian Pacific Journal of Tropical Medicine 17, 191 (2024).

34. Wong, L. P., Shakir, S. M. M., Atefi, N. & AbuBakar, S. Factors Affecting Dengue Prevention Practices: Nationwide Survey of the Malaysian Public. PLOS ONE 10, e0122890 (2015).

35. Duval, P., Aschan-Leygonie, C. & Valiente Moro, C. A review of knowledge, attitudes and practices regarding mosquitoes and mosquito-borne infectious diseases in nonendemic regions. Front. Public Health 11, (2023).

36. Caputo, B., Manica, M., Russo, G. & Solimini, A. Knowledge, Attitude and Practices towards the Tiger Mosquito Aedes Albopictus. A Questionnaire Based Survey in Lazio Region (Italy) before the 2017 Chikungunya Outbreak. International Journal of Environmental Research and Public Health 17, 3960 (2020).

37. Molho, S. et al. Public perceptions and prevention behaviours related to arboviral diseases in Metropolitan France: 2016 Health Barometer. Bulletin Epidémiologique Hebdomadaire, 24, 510–517 (2018).

38. Constant, A., McColl, K. & Raude, J. The Ecology of Protective Behaviors: A Study in New Risk Areas for Mosquito-Borne Diseases. EcoHealth 17, 315–325 (2020).

39. Raude, J. et al. Public perceptions and behaviours related to the risk of infection with Aedes mosquito-borne diseases: a cross-sectional study in Southeastern France. BMJ Open 2, e002094 (2012).

40. de Best, P. A. et al. Determinants of intended prevention behaviour against mosquitoes and mosquito-borne viruses in the Netherlands and Spain using the MosquitoWise survey: cross-sectional study. BMC Public Health 24, 1781 (2024).

41. Duval, P., Valiente Moro, C. & Aschan-Leygonie, C. How do attitudes shape protective practices against the Asian tiger mosquito in community gardens in a nonendemic country? Parasites Vectors 15, 439 (2022).

42. Le Tyrant, M., Bley, D., Leport, C., Alfandari, S. & Guégan, J.-F. Low to medium-low risk perception for dengue, chikungunya and Zika outbreaks by infectious diseases physicians in France, Western Europe. BMC Public Health 19, 1014 (2019).

43. Farooq, Z. et al. Impact of climate and Aedes albopictus establishment on dengue and chikungunya outbreaks in Europe: a time-to-event analysis. The Lancet Planetary Health 9, e374–e383 (2025).

44. McNeil, C., Verlander, S., Divi, N. & Smolinski, M. The Landscape of Participatory Surveillance Systems Across the One Health Spectrum: Systematic Review. JMIR Public Health and Surveillance 8, e38551 (2022).

45. Guerrisi, C. et al. Participatory Syndromic Surveillance of Influenza in Europe. The Journal of Infectious Diseases 214, S386–S392 (2016).

46. De Meijere, G. et al. Attitudes towards booster, testing and isolation, and their impact on COVID-19 response in winter 2022/2023 in France, Belgium, and Italy: a cross-sectional survey and modelling study. The Lancet Regional Health - Europe 28, 100614 (2023).

47. Kelley, K., Gozzi, N., Mazzoli, M. & Paolotti, D. Exploring influenza vaccination determinants through digital participatory surveillance. BMC Public Health 25, 1345 (2025).

48. Gozzi, N., Perrotta, D., Paolotti, D. & Perra, N. Towards a data-driven characterization of behavioral changes induced by the seasonal flu. PLOS Computational Biology 16, e1007879 (2020).

49. McColl, K. et al. Are People Optimistically Biased about the Risk of COVID-19 Infection? Lessons from the First Wave of the Pandemic in Europe. IJERPH 19, 436 (2021).

50. Debin, M. et al. Evaluating the Feasibility and Participants’ Representativeness of an Online Nationwide Surveillance System for Influenza in France. PLoS ONE 8, e73675 (2013).

51. FOR2 - Population non scolarisée de 15 ans ou plus par sexe, âge et diplôme le plus élevé en 2019 − France entière −Diplômes - Formation en 2019 | Insee. (2019). https://www.insee.fr/fr/statistiques/6455250?sommaire=6455252&geo=FE-1 [online, accessed: 05/11/2025]

52. Grado di istruzione dettagliato della popolazione residente di 6 anni e più. (2022).

53. Rodríguez, G. Chapter 6: Multinomial Response Models. Preprint at https://grodri.github.io/glms/notes/ (2007).

54. Bürkner, P.-C. & Vuorre, M. Ordinal Regression Models in Psychology: A Tutorial. Advances in Methods and Practices in Psychological Science 2, 77–101 (2019).

55. Liddell, T. M. & Kruschke, J. K. Analyzing ordinal data with metric models: What could possibly go wrong? Journal of Experimental Social Psychology 79, 328–348 (2018).

56. Harrell, F. E. Regression Modeling Strategies: With Applications to Linear Models, Logistic and Ordinal Regression, and Survival Analysis. (Springer International Publishing, Cham, 2015). doi:10.1007/978-3-319-19425-7.

57. Ordinal Logistic Regression| R data analysis examples. UCLA: Statistical Consulting Group. https://stats.oarc.ucla.edu/r/dae/ordinal-logistic-regression/. [online, accessed: 05/11/2025]

58. Principali statistiche geografiche sui comuni – Istat. https://www.istat.it/classificazione/principali-statistiche-geografiche-sui-comuni/. [online, accessed: 05/11/2025]

59. Une nouvelle définition du rural pour mieux rendre compte des réalités des territoires et de leurs transformations − La France et ses territoires | Insee. https://www.insee.fr/fr/statistiques/5039991?sommaire=5040030. [online, accessed: 05/11/2025]

60. Chikungunya, dengue et zika - Données de la surveillance renforcée en France métropolitaine en 2023. https://www.santepubliquefrance.fr/maladies-et-traumatismes/maladies-a-transmission-vectorielle/chikungunya/articles/donnees-en-france-metropolitaine/chikungunya-dengue-et-zika-donnees-de-la-surveillance-renforcee-en-france-metropolitaine-en-2023. [online, accessed: 05/11/2025]

61. EpiCentro. Febbre dengue - News. https://www.epicentro.iss.it/febbre-dengue/2021-2023. [online, accessed: 05/11/2025]

62. Dengue. European Centre for Disease Prevention and Control https://www.ecdc.europa.eu/en/dengue (2010). [online, accessed: 05/11/2025]

63. Guerrisi, C. et al. Factors associated with influenza-like-illness: a crowdsourced cohort study from 2012/13 to 2017/18. BMC Public Health 19, 879 (2019).

64. Liard, R. et al. Seasonal influenza vaccination in pharmacy in France: description and determinants of the vaccinated at-risk population using this service, 1 year after implementation. International Journal of Pharmacy Practice 30, 253–260 (2022).

65. Cipolletta, S., Andreghetti, G. & Mioni, G. Risk Perception towards COVID-19: A Systematic Review and Qualitative Synthesis. IJERPH 19, 4649 (2022).

66. Clark, C., Davila, A., Regis, M. & Kraus, S. Predictors of COVID-19 voluntary compliance behaviors: An international investigation. Global Transitions 2, 76–82 (2020).

67. Wachira, E., Laki, K., Chavan, B., Aidoo-Frimpong, G. & Kingori, C. Factors Influencing COVID-19 Prevention Behaviors. J of Prevention 44, 35–52 (2023).

68. Seale, H. et al. COVID-19 is rapidly changing: Examining public perceptions and behaviors in response to this evolving pandemic. PLoS ONE 15, e0235112 (2020).

69. Tizzani, M. & Gauvin, L. Socioeconomic determinants of protective behaviors and contact patterns in the post-COVID-19 pandemic era: A cross-sectional study in Italy. PLOS Computational Biology 21, e1013262 (2025).

70. Callaghan, T., Lueck, J. A., Trujillo, K. L. & Ferdinand, A. O. Rural and Urban Differences in COVID-19 Prevention Behaviors. The Journal of Rural Health 37, 287–295 (2021).

71. Kraemer, M. U. G. et al. The global compendium of Aedes aegypti and Ae. albopictus occurrence. Sci Data 2, 150035 (2015).

72. Wilke, A. B. B. et al. Urbanization creates diverse aquatic habitats for immature mosquitoes in urban areas. Sci Rep 9, 15335 (2019).

73. Neumann-Böhme, S. et al. Once we have it, will we use it? A European survey on willingness to be vaccinated against COVID-19. Eur J Health Econ 21, 977–982 (2020).

74. Bish, A., Yardley, L., Nicoll, A. & Michie, S. Factors associated with uptake of vaccination against pandemic influenza: A systematic review. Vaccine 29, 6472–6484 (2011).

75. Zintel, S. et al. Gender differences in the intention to get vaccinated against COVID-19: a systematic review and meta-analysis. J Public Health (Berl.) 31, 1303–1327 (2023).

76. Annan, E., Treviño, J., Zhao, B., Rodriguez-Morales, A. J. & Haque, U. Direct and indirect effects of age on dengue severity: The mediating role of secondary infection. PLoS Negl Trop Dis 17, e0011537 (2023).

77. Liang, B. & Scammon, D. L. Incidence of Online Health Information Search: A Useful Proxy for Public Health Risk Perception. J Med Internet Res 15, e114 (2013).

78. van Noort, S. P. et al. Ten-year performance of Influenzanet: ILI time series, risks, vaccine effects, and care-seeking behaviour. Epidemics 13, 28–36 (2015).

79. Cantarelli, P. et al. The representativeness of a European multi-center network for influenza-like-illness participatory surveillance. BMC Public Health 14, 984 (2014).

